# Healthcare coverage and associated factors in Cameroon: analyses from a national survey

**DOI:** 10.1101/2025.06.22.25330087

**Authors:** Fabrice Zobel Lekeumo Cheuyem, Constantine Tanywe Asahngwa, Solange Dabou, Brian Ngongheh Ajong, Guy Stéphane Nloga, Jessy Goupeyou-Youmsi, Ariane Nouko, Edwige Omona Guissana, Rick Tchamani, Ethel Ambo Eno, Achangwa Chabeja, Innocent Takougang

**Affiliations:** Department of Public Health, Faculty of Medicine and Biomedical Sciences, The University of Yaoundé 1, Yaoundé, Cameroon; Division of Health Policy and Research, Nkafu Policy Institute, Denis and Lenora Foretia Foundation, Yaoundé, Cameroon; Department of Anthropology, The University of Yaoundé 1, Yaoundé, Cameroon; Health Emergencies Program, World Health Organization (WHO), Kinshasa, Democratic Republic of Congo; Department of Family Health, The Ministry of Public Health, Yaoundé, Cameroon; Rosière Higher School of Health Sciences, Yaoundé, Cameroon; Women in Global Health, Yaounde, Cameroon; Department of Public Health, University of West Indies, Bridgetown, Barbados

**Keywords:** Health insurance, universal health coverage, medical coverage, health coverage, healthcare access, out-of-pocket payments, Cameroon

## Abstract

**Background:** Emergency medicine systems are vital for reducing mortality and disability, yet Cameroon faces significant healthcare access challenges, with only 0.6% of GDP allocated to public health and 70% of health expenditures paid out-of-pocket. This study assessed proportion of population benefiting from health coverage, patterns, and associated factors in Cameroon to inform policies aimed at achieving universal health coverage (UHC).

**Methods:** A nationally representative cross-sectional survey was conducted from February to March 2024, involving 1,200 adults selected via multistage random sampling. Data were collected through face-to-face interviews by national Afrobarometer team, and analyzed with R Statistics version 4.4.2. Logistic regression identified factors associated with medical coverage, adjusting for sociodemographic, economic, and occupational variables.

**Results:** A proportion of 7.9% (95% CI: 6.4–9.6) of respondents reported benefiting from a health coverage in 2024, with the lowest rates reported in the North, Adamawa, Centre, and North-West regions. Private health insurance was the most common (44.3%), followed by civil servant schemes (20.5%) and community-based insurance (15.9%). Unemployed individuals were twice as likely to lack coverage (aOR = 2.22, 95% CI: 1.42–3.51). Those with secondary education had twice the odds of being uninsured compared to tertiary-educated individuals (aOR = 2.00, 95% CI: 1.21–3.27). Insured individuals were more likely to use healthcare services (9.6%; p = 0.028), reported easier access to medical care (15.8%; p = 0.010), and expressed fewer concerns about healthcare access (23.4%; p < 0.001). Barriers among the uninsured included high costs (37.1%), lack of information (31.6%), complex registration processes (8.5%) and geographical constraints. Most of the community members reported being quite (51.1%) or very satisfied (23.9%) by the medical coverage they are benefiting from. Notably, 68% of respondents supported higher taxes to improve healthcare access.

**Conclusion:** Cameroon’s suboptimal medical coverage reflects systemic inequities tied to employment and education. Expanding employer-independent schemes, subsidizing premiums, and leveraging community-based models are critical to advancing UHC. Public willingness to contribute through taxes suggests political viability for systemic reforms.

## Background

Emergency medicine systems are key players in the global healthcare architecture, saving lives and minimizing long-term disabilities. Their core function is to deliver urgent, time-critical medical care designed to significantly improve a patient’s chances of recovery without lasting impairment [1,2]. It culminates in the immediate provision of life-saving emergency care upon arrival at the hospital, often within hours, minutes mere seconds, or of symptom onset [3]. Globally there is still a high proportion of people dying in emergency department of many healthcare settings. In 2019, emergency and/or operative conditions accounted for 514.09 deaths per 100,000 population and 18,113.00 Disability-adjusted life year (DALYs) per 100,000 population worldwide [4]. For emergency conditions, the per capita burden of deaths and DALYs is greatest for low-income countries [5].

In Cameroon, the life expectancy at birth (61 years in 2020) is still lower than that of several developed countries [6]. The maternal mortality ratio in 2023 is 250/100,000 live births, reflecting a high risk of women dying while giving birth [7]. A significant proportion of these deaths occur in emergency situations such as hemorrhage due to road traffic accidents or post-partum hemorrhage for women [8,9]. To properly address such health emergencies, a rapid pooling of health human resources, an adequate and high-quality technical platform, and financial resources are required. In a context of limited resources, mobilizing these resources for some families to save the life of their relatives might be challenging to ensure the optimal management of the patient.

In countries like Cameroon, domestic public health spending, at a mere 0.6% of gross domestic product, ranks as the third-lowest in sub-Saharan Africa [10]. Cameroon also faces the fourth-highest household out-of-pocket expenditure in the region, with approximately 475 billion XAF annually, representing 70% of total health expenditure [10,11]. In this context, where roughly 4 in 10 Cameroonians live below the national poverty line, health issues, especially emergency situations, might lead to death due to delays in patient management [12]. Additionally, the utilization of healthcare services (0.41 per 10,000 population) is still very low compared to the WHO target (5 per 10,000 population). This leads to delayed seeking care during illness, which worsens the patient’s condition and reduces the chances of recovery [6].

To prevent these deaths resulting from limited financial resources and poor utilization of healthcare services, the implementation of universal health coverage (UHC) in the country is one possible solution to increase access to quality healthcare [13,14]. Since April 2022, Cameroon has engaged in a significant UHC health system reform, aiming to ensure fair access to high-quality, affordable healthcare for all its citizens, with a special focus on the nation’s most vulnerable groups [15].

However, the packages offered in this initial phase primarily covered healthcare for specific groups within the Cameroonian population [16].This package includes several interventions ranging from prevention to care and treatment services for pregnant women, children under 5 years, patients on dialysis, and those diagnosed with tuberculosis, as well as human immunodeficiency virus (HIV)-related health services for people living with HIV [17].

In order to benefit from medical coverage, other population groups not covered by this UHC must subscribe to one of the health insurance schemes available in the country. Such medical insurance options include community-based health insurance, private health insurance offered by some traditional insurance and bank companies, and other medical coverages offered by the government to civil servants [14,18].

To sustainably run the UHC public policy in the country, households will be required to contribute financially to the program. Study reports identified significant factors associated with willingness to pay for UHC. Such factors include sociodemographic and economic characteristics, healthcare utilization, and policy perception [15].

On the other hand, a study conducted in Cameroon revealed six primary barriers to health insurance uptake and utilization, affecting both subscribers and non-subscribers. These included distrust in current health insurance policies, insufficient awareness of available schemes, high subscription costs, lengthy and complex bureaucratic procedures, infrequent healthcare utilization, and the common practice of self-medication [14].

To better inform decision-makers, there’s a clear need for data on the implementation of healthcare coverage in Cameroon as perceived by the general population. This information would help national and international stakeholders to develop targeted strategies to improve access to quality healthcare, reduce out-of-pocket (OOP) expenses, and prevent avoidable deaths due to financial constraints. This study aimed to provide a comprehensive national overview of how healthcare coverage is implemented in Cameroon. Specifically, we need to map the distribution of medical coverage across the country, identify the factors that determine its uptake, reveal the most prevalent types of insurance, understand the behaviors of both insured and uninsured individuals, and assess the level of satisfaction among beneficiaries.

## Methods

### Study design and period

The present study was cross-sectional survey conducted in Cameroon from February 27 and March 20, 2024.

### Study setting

Cameroon, located in Central Africa, is a geographically and culturally diverse nation often referred to as ‘Africa in miniature’ due to its varied landscapes, climates, and ethnic groups, with French and English as official languages [19]. Bordered by Nigeria, Chad, the Central African Republic, Equatorial Guinea, Gabon, and the Republic of the Congo, it features coastal plains, dense rainforests, savannas, and mountainous regions [19,20]. The country has an estimated population of over 29 million people, it is divided in ten administrative regions; it has two capital cities: Yaoundé, the political capital, and Douala, the economic capital [21]. The population is very young: 43.6% of Cameroonians are less than 15 years [22]. Cameroon’s economy is mixed, relying on agriculture, oil, timber, and mining, while its political landscape has been dominated by long-standing stability, though recent years have seen tensions in Anglophone regions (South-West and North-West) [23].

### Study participants and sampling

This survey included adult Cameroonians aged 18 and more who provided their informed consent to participate. A multistage random sampling technique was used to select a total of 1,200 nationally representative adult Cameroonians to yield national results with a margin of error of +/-3 percentage points at a 95% confidence level [24].

### Data collection

Data collection was performed by the Afrobarometer Cameroon national partner, led by Cible Etudes & Conseil. Afrobarometer is a pan-African, non-partisan survey research network that gathers reliable data on Africans’ experiences and evaluations of democracy, governance, and quality of life. The interview was conducted face-to-face in the respondent’s language with nationally representative samples. Afrobarometer provides support and oversight at each stage of the process [25].

National partners trained interviewers to ensure that they are familiar with the questionnaire in national and local languages, the sampling protocol, and best field practices, and are confident in applying Afrobarometer’s survey methodology. After the training, teams of four interviewers and one field supervisor travelled to enumeration areas in all parts of the country. Each national sample was distributed across regions and urban/rural areas in proportion to their share in the national population. Every adult citizen had an equal chance of being selected as a respondent. Interviews were conducted in a language of the respondent’s choice using electronic tablets, and usually took about an hour. Responses were strictly confidential and anonymized [25].

### Data quality assessment

The national partner verified and checked their data for any incomplete, improperly formatted, or inaccurate records. Data managers at regional core partners and at Afrobarometer cleaned and finalized the data sets. A more detailed methodology used to implement this survey is available online [25].

### Data source and statistical analysis

Data were downloaded from the online repository of Afrobarometer [26]. They were then exported, recoded as necessary, and analyzed using R Statistics version 4..4.2 [27]. The respondent characteristics were presented as counts and frequencies. The Chi-2 and Fisher exact tests were used to establish associations between categorical variables. Maps were generated using QGIS Desktop software version 3.42.3. The dependent variable was benefiting from any form a medical coverage, while the independent variables included age, sex, educational level, religion, economic status, occupation status (having a job or not), and professional status. Univariable and multivariable logistic regression analyses were used to identify factors associated with medical coverage through Odds Ratios (ORs) and their corresponding 95% confidence intervals (CIs). The selection of predictors that best fit the model was done stepwise using the Akaike Information Criterion (AIC) [28]. The model with the lowest index was then selected [29]. A *p*-value ⍰5% was considered statistically significant.

## Results

A total of 1,200 Cameroonians were enrolled in this study. Most of them were aged less than 36 years (68%), male and female as well as rural and urban residents were equally represented. They mostly attended secondary school (53%) (Table 1).

**Table 1.** Sociodemographic and professional characteristics of study participants, 2024 (n = 1,200)

| Characteristic | Count (n) | Frequency (%) |
| --- | --- | --- |
| **Age group** |  |  |
| 18-25 | 376 | 31.3 |
| 26-35 | 438 | 36.5 |
| 36-45 | 216 | 18.0 |
| 46-55 | 108 | 9.0 |
| 56+ | 62 | 5.2 |
| **Sex** |  |  |
| Female | 595 | 49.6 |
| Male | 605 | 50.4 |
| **Residence area** |  |  |
| Rural | 592 | 49.3 |
| Urban | 608 | 50.7 |
| **Educational level** |  |  |
| No formal education | 88 | 7.3 |
| Primary | 187 | 15.6 |
| Secondary | 641 | 53.4 |
| Tertiary | 284 | 23.7 |
| **Region of residence** |  |  |
| Adamawa | 56 | 4.7 |
| Centre | 248 | 20.7 |
| East | 56 | 4.7 |
| Far-North | 184 | 15.3 |
| Littoral | 216 | 18.0 |
| North | 128 | 10.7 |
| North-West | 104 | 8.7 |
| South | 40 | 3.3 |
| South-West | 96 | 8.0 |
| West | 72 | 6.0 |
| **Religion** |  |  |
| Christian | 925 | 77.1 |
| Muslim | 203 | 16.9 |
| Other | 72 | 6.0 |
| **Main occupation** |  |  |
| Agro-pastoral work | 213 | 17.8 |
| Skilled professionals | 207 | 17.3 |
| Housewife | 111 | 9.3 |
| Middle-ranking professional | 115 | 9.6 |
| Never had a job | 40 | 3.3 |
| Merchant | 251 | 20.9 |
| Student | 182 | 15.2 |
| Unskilled worker | 81 | 6.8 |
| **Perceived economic situation** |  |  |
| Very bad | 247 | 20.6 |
| Bad | 311 | 25.9 |
| Fair | 229 | 19.1 |
| Fairly good | 345 | 28.7 |
| Very good | 68 | 5.7 |

**Table 2.** Community member perceptions on healthcare service according to their reported health coverage status in Cameroon, 2024 (n = 1,200)

| Variable | Health coverage |  | Total | *p*-value<sup>†</sup> |
| --- | --- | --- | --- | --- |
|  | Yes<br>n (%) | No<br>n (%) |  |  |
|  | N = 88 | N = 1,112 | 1,200 (100) |  |
| **Dealt with a health facility** |  |  |  | 0.028 |
| No | 47 (6.1) | 725 (93.9) | 772 |  |
| Yes | 41 (9.6) | 387 (90.4) | 428 |  |
| **Obtaining medical attention** |  |  |  | 0.010 |
| Very difficult | 5 (8.6) | 53 (91.4) | 58 |  |
| Difficult | 8 (5.2) | 146 (94.8) | 154 |  |
| Easy | 22 (12.4) | 155 (87.6) | 177 |  |
| Very easy | 6 (15.8) | 32 (84.2) | 38 |  |
| No contact/Non applicable | 47 (6.1) | 726 (93.9) | 773 |  |
| **Concerns about healthcare access** |  |  |  | <0.001 |
| Very much | 21 (3.6) | 570 (96.4) | 591 |  |
| A little | 19 (9.0) | 192 (91.0) | 211 |  |
| Somewhat | 23 (7.9) | 268 (92.1) | 291 |  |
| Not at all | 25 (23.4) | 82 (76.6) | 107 |  |
| **More taxes to guarantee access to healthcare** |  |  |  | 0.668 |
| Strongly agree | 31 (7.4) | 387 (92.6) | 418 |  |
| Agree | 25 (6.3) | 371 (93.7) | 396 |  |
| Neutral | 7 (11.3) | 55 (88.7) | 62 |  |
| Disagree | 16 (7.8) | 188 (92.2) | 204 |  |
| Strongly disagree | 9 (7.5) | 111 (92.5) | 120 |  |
| **Frequency of lack of healthcare** |  |  |  | 0.791 |
| Never | 25 (8.0) | 286 (92.0) | 311 |  |
| Just one or two times | 21 (8.6) | 222 (91.4) | 243 |  |
| sometimes | 21 (6.1) | 324 (93.9) | 345 |  |
| Many times | 16 (7.1) | 209 (92.9) | 225 |  |
| Always | 5 (6.6) | 71 (93.4) | 76 |  |

<sup>†</sup> Pearson's chi square test

### Health coverage pattern

A low proportion of Cameroonian (7.9%; 95% CI: 6.4-9.6) reported benefiting from a health coverage. The regions whose residents reported the lowest health coverage included the North, the Adamawa, the Centre, and the North-West. The subdivision with the lowest reported health coverage encompassed the Dja-et-Lobo in the South region, the Faro and the Mayo-Louti in the North region, the Mayo-Banyo and the Mbéré in the Adamawa region, the Mbam-et-Inoubou and the Nyong-et-So’o in the Centre region, and the Mifi and the Nkam in the West region (Fig. 1 and Supplementary Table 1 and 2).

**Fig. 1.**
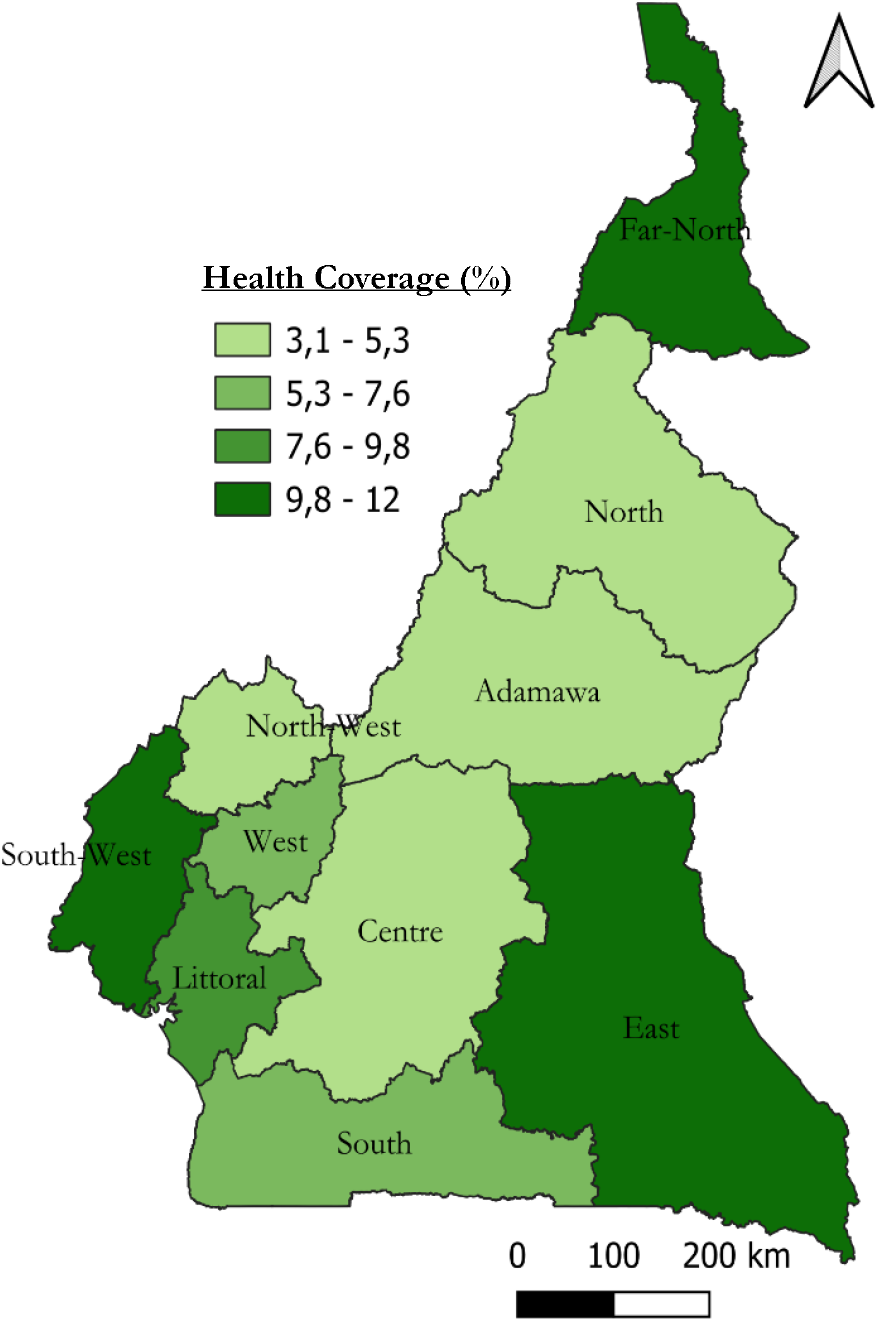
Regional mapping of health coverage among community members in Cameroon, 2024.

There was a significant association between being covered by health insurance and using the healthcare service in the last 12 months (9.6%; *p* = 0.028). Community members covered tended to significantly perceive the obtention of medical attention very easy (15.8%; *p* = 0.010). Furthermore, those who benefited from health coverage reported less concerns about healthcare access (23.4%; *p* ⍰0.001). Moreover, more than two-thirds of the respondents (n = 814; 68%) strongly agree or just agree to pay more taxes to warrant better access to healthcare services (Table 2).

The most commonly reported health insurance benefited included the private health insurance (44.3%), civil servant insurance scheme (20.5%) and the community-based health insurance (15.9%) (Fig 2).

**Fig. 2.**
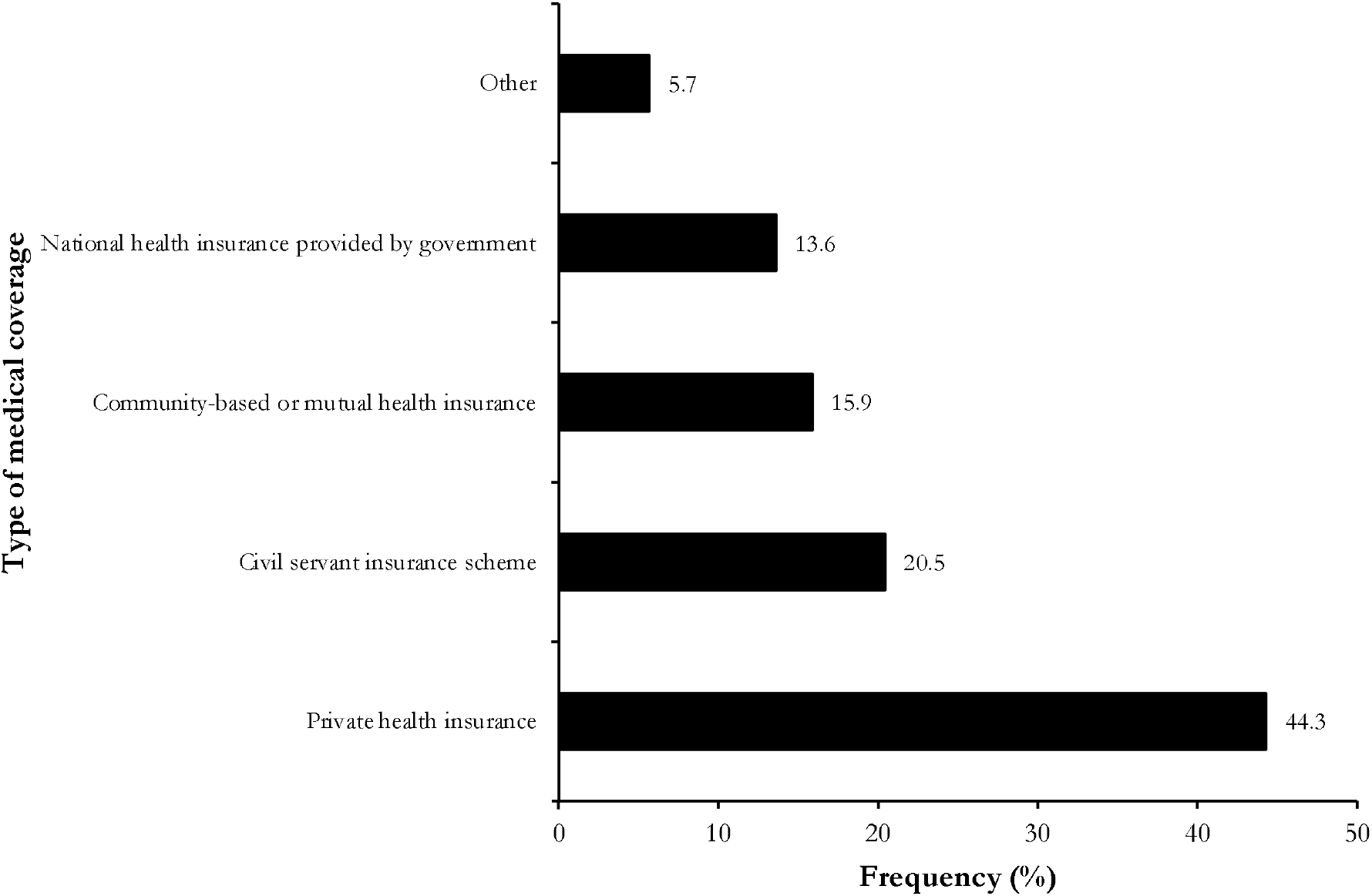
Reported patterns of health coverages benefited by Cameroonian, 2024 (n = 88)

Most of the community members reported being quite (51.1%) or very satisfied (23.9%) by the health coverage they are benefiting from (Fig 3).

**Fig. 3.**
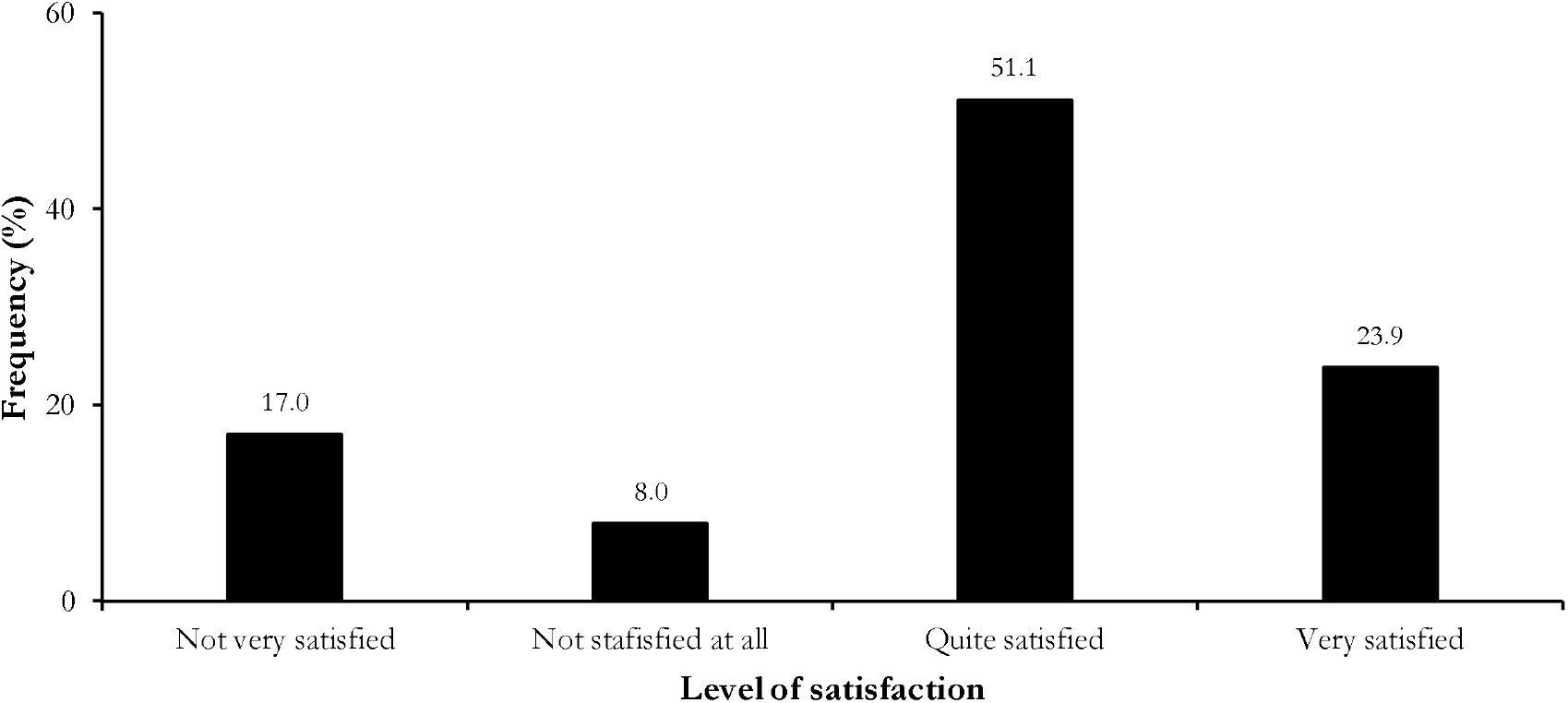
Level of satisfaction about the health coverage among beneficiaries in Cameroon, (n = 88)

Among 1,112 who reported not benefiting from any health coverage, the most commonly reported barrier included the cost which was judged high (37.1%), lack of information related to health insurance schemes (31.6%), registration perceived as complex (8.5%), and geographical constraints (7.1%) (Fig. 4).

**Fig. 4.**
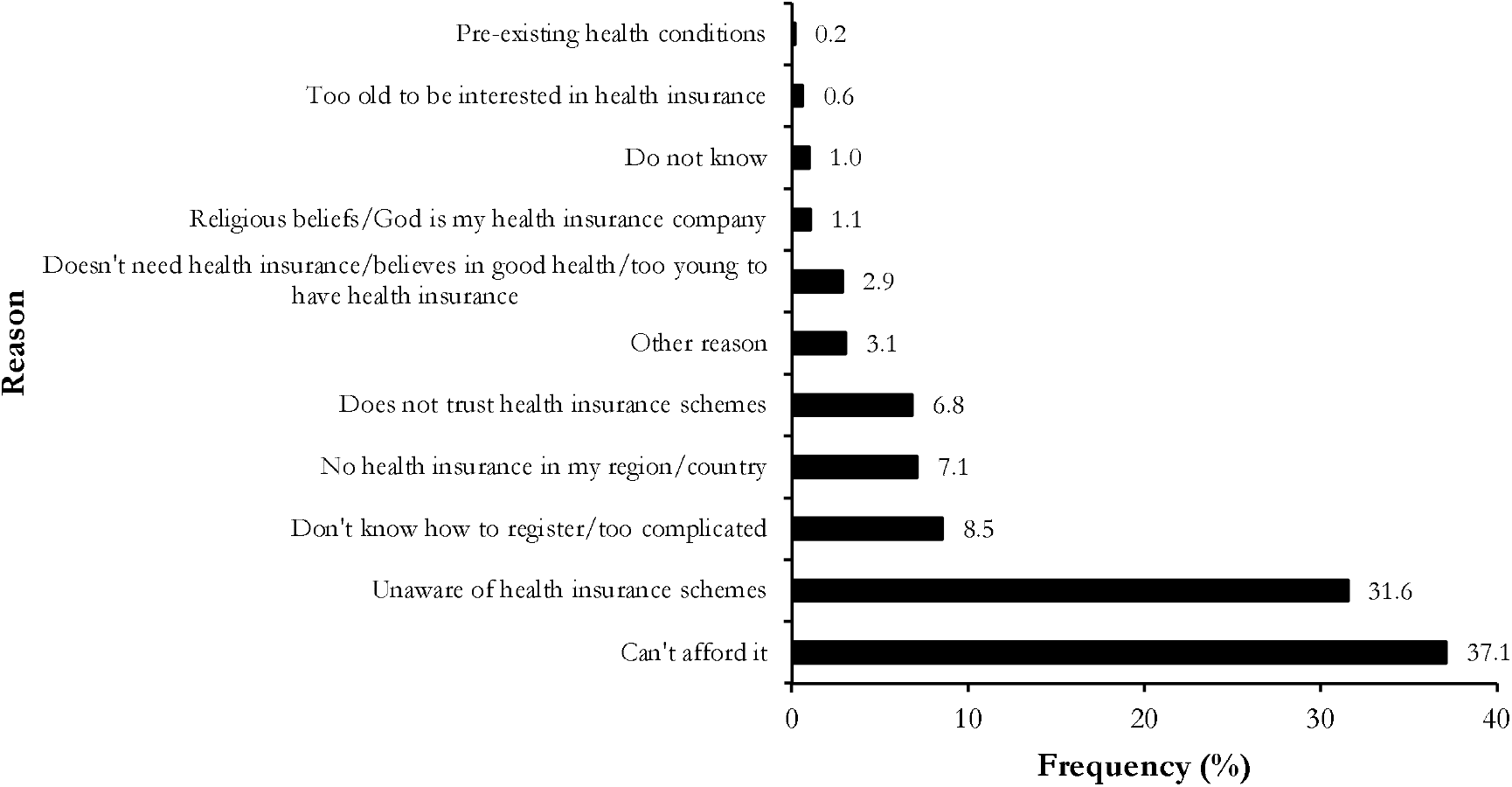
Reported barrier to health coverage access among Cameroonians, 2024 (n = 1,112)

### Factors associated with health coverage

Based on the multivariate analysis, benefiting from health coverage were significantly associated with owning a job (*p* ⍰0.001) and community member’s educational level (*p* = 0.006). Those who had no job in the last twelve months were significantly twice more likely to report being uncovered by a health insurance (aOR = 2.22; 95% CI: 1.42-3.51). The lesser the education level, the higher the odds of being uncovered; compared to those with tertiary level of education, community members with secondary level of education were significantly twice (aOR = 2.00; 95% CI: 1.21-3.27) more likely to be uncovered by a health insurance scheme (Table 3).

**Table 3.**
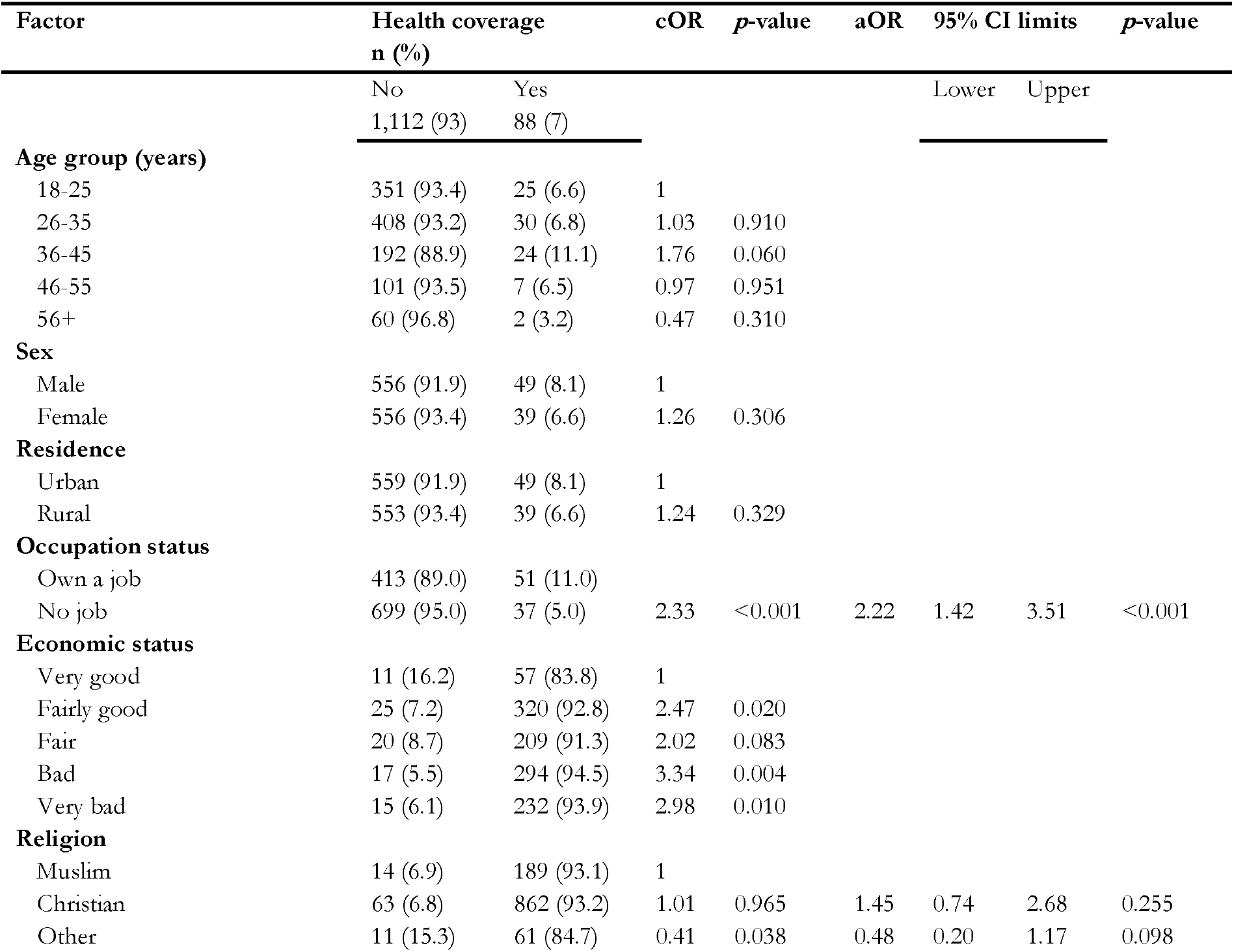
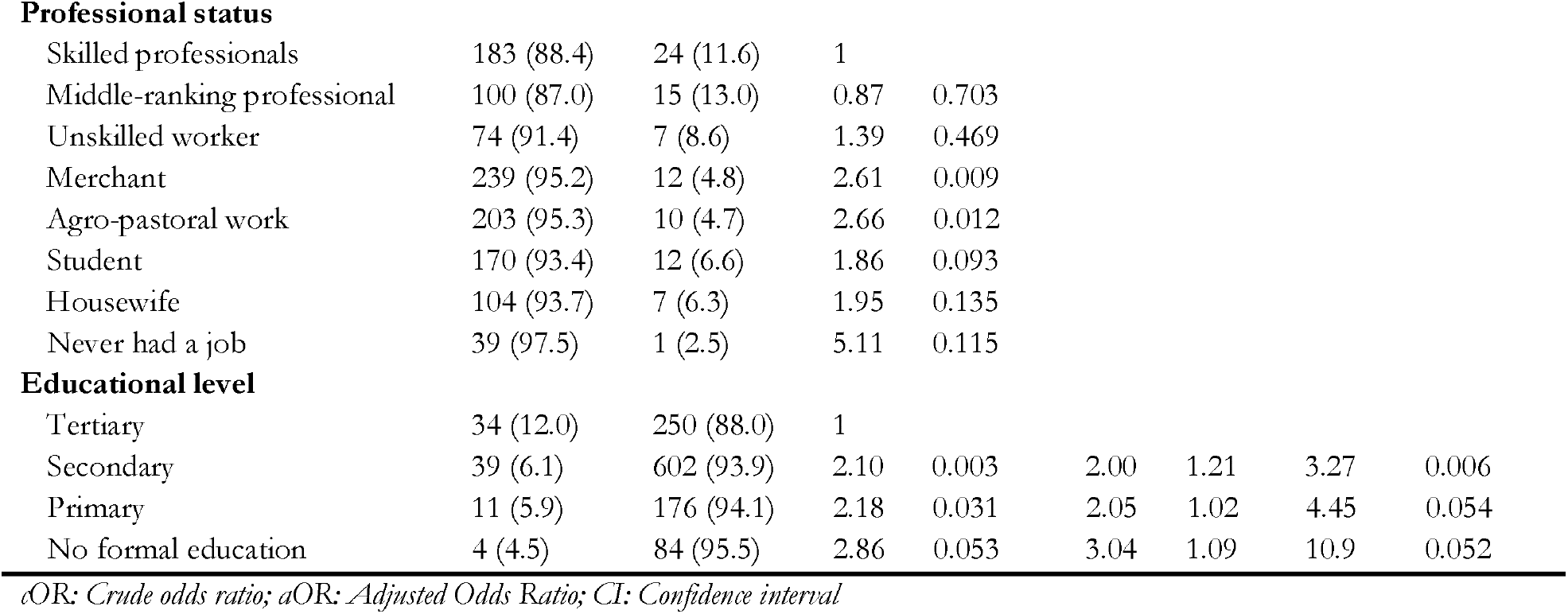
Simple and multiple binary logistic regressions of socioprofessional and economic parameters associated with non-coverage by health insurance among Cameroonians, 2024 (n = 1,200)

| Factor | Health coverage<br>n (%) |  | cOR | p-value | aOR | 95% CI limits |  | p-value |
| --- | --- | --- | --- | --- | --- | --- | --- | --- |
|  | No | Yes |  |  |  | Lower | Upper |  |
|  | 1,112 (93) | 88 (7) |  |  |  |  |  |  |
| **Age group (years)** |  |  |  |  |  |  |  |  |
| 18-25 | 351 (93.4) | 25 (6.6) | 1 |  |  |  |  |  |
| 26-35 | 408 (93.2) | 30 (6.8) | 1.03 | 0.910 |  |  |  |  |
| 36-45 | 192 (88.9) | 24 (11.1) | 1.76 | 0.060 |  |  |  |  |
| 46-55 | 101 (93.5) | 7 (6.5) | 0.97 | 0.951 |  |  |  |  |
| 56+ | 60 (96.8) | 2 (3.2) | 0.47 | 0.310 |  |  |  |  |
| **Sex** |  |  |  |  |  |  |  |  |
| Male | 556 (91.9) | 49 (8.1) | 1 |  |  |  |  |  |
| Female | 556 (93.4) | 39 (6.6) | 1.26 | 0.306 |  |  |  |  |
| **Residence** |  |  |  |  |  |  |  |  |
| Urban | 559 (91.9) | 49 (8.1) | 1 |  |  |  |  |  |
| Rural | 553 (93.4) | 39 (6.6) | 1.24 | 0.329 |  |  |  |  |
| **Occupation status** |  |  |  |  |  |  |  |  |
| Own a job | 413 (89.0) | 51 (11.0) |  |  |  |  |  |  |
| No job | 699 (95.0) | 37 (5.0) | 2.33 | <0.001 | 2.22 | 1.42 | 3.51 | <0.001 |
| **Economic status** |  |  |  |  |  |  |  |  |
| Very good | 11 (16.2) | 57 (83.8) | 1 |  |  |  |  |  |
| Fairly good | 25 (7.2) | 320 (92.8) | 2.47 | 0.020 |  |  |  |  |
| Fair | 20 (8.7) | 209 (91.3) | 2.02 | 0.083 |  |  |  |  |
| Bad | 17 (5.5) | 294 (94.5) | 3.34 | 0.004 |  |  |  |  |
| Very bad | 15 (6.1) | 232 (93.9) | 2.98 | 0.010 |  |  |  |  |
| **Religion** |  |  |  |  |  |  |  |  |
| Muslim | 14 (6.9) | 189 (93.1) | 1 |  |  |  |  |  |
| Christian | 63 (6.8) | 862 (93.2) | 1.01 | 0.965 | 1.45 | 0.74 | 2.68 | 0.255 |
| Other | 11 (15.3) | 61 (84.7) | 0.41 | 0.038 | 0.48 | 0.20 | 1.17 | 0.098 |

**Professional status**
|  |  |  |  |  |
| --- | --- | --- | --- | --- |
| Skilled professionals | 183 (88.4) | 24 (11.6) | 1 |  |
| Middle-ranking professional | 100 (87.0) | 15 (13.0) | 0.87 | 0.703 |
| Unskilled worker | 74 (91.4) | 7 (8.6) | 1.39 | 0.469 |
| Merchant | 239 (95.2) | 12 (4.8) | 2.61 | 0.009 |
| Agro-pastoral work | 203 (95.3) | 10 (4.7) | 2.66 | 0.012 |
| Student | 170 (93.4) | 12 (6.6) | 1.86 | 0.093 |
| Housewife | 104 (93.7) | 7 (6.3) | 1.95 | 0.135 |
| Never had a job | 39 (97.5) | 1 (2.5) | 5.11 | 0.115 |
| **Educational level** |  |  |  |  |  |  |  |  |
| Tertiary | 34 (12.0) | 250 (88.0) | 1 |  |  |  |  |  |
| Secondary | 39 (6.1) | 602 (93.9) | 2.10 | 0.003 | 2.00 | 1.21 | 3.27 | 0.006 |
| Primary | 11 (5.9) | 176 (94.1) | 2.18 | 0.031 | 2.05 | 1.02 | 4.45 | 0.054 |
| No formal education | 4 (4.5) | 84 (95.5) | 2.86 | 0.053 | 3.04 | 1.09 | 10.9 | 0.052 |

*cOR: Crude odds ratio; aOR: Adjusted Odds Ratio; CI: Confidence interval*

## Discussion

This study reveals a low proportion (7.9%) of Cameroonians benefiting from any form of health coverage, highlighting significant gaps in healthcare access across the country. The disparities in coverage observed, align with broader socio-demographic, economic and geographic inequalities, where rural and economically disadvantaged populations face greater barriers to healthcare services [30–32]. Our result corroborates findings from other previous survey conducted in 2020 in Cameroon and in Sub-Saharan Africa [33]. This highlights the persistence of a low health insurance coverage within communities across time. However, the progressive increase in the basket of care and the modalities allowing the inclusion of a greater number of beneficiaries through the implementation of the next phases of UHC in Cameroon will make it possible to improve the health coverage of the population and prevent catastrophic out-of-pocket expenses for health. Furthermore, studies have reported recurrent hospital detentions due to unpaid hospital bills among the most vulnerable strata of the Cameroonian population, highlighting the need for a functional healthcare protection mechanism to reduce the cost of healthcare services [34,35]. This low healthcare coverage in Cameroon underscores the urgent need for policy interventions to expand health insurance especially the UHC and improve healthcare accessibility, particularly in underserved regions such as the North, Adamawa, Centre, and North-West

The study identified key factors associated with health coverage, including employment status and educational attainment. Individuals without jobs were twice as likely to lack health insurance (aOR = 2.22), emphasizing the role of formal employment in facilitating access to coverage, likely through employer-sponsored schemes or through individual direct subscription to a private health insurance scheme [36]. Similarly, lower educational levels were strongly correlated with non-coverage, with those having only secondary education being twice as likely to be uninsured compared to those with tertiary education (aOR = 2.00). These findings suggest that education may enhance awareness of health insurance benefits and the ability to navigate enrollment processes; as the lengthy and complicated bureaucratic processes has been reported as one of the barrier to health insurance subscription in Cameroon [14]. Furthermore, in addition to better awareness, the most educated individuals might perceive better the usefulness of healthcare coverage and trust health insurance companies more. Our findings confirm observations from a multilevel national survey in sub-Saharan Africa [33].

The type of health insurance varied among beneficiaries, with private health insurance being the most common (44.3%), followed by civil servant schemes (20.5%) and community-based insurance (15.9%). This distribution reflects the fact that health insurance system is often tied to employment or individual ability to purchase an insurance scheme, leaving vulnerable groups such as informal workers and rural residents disproportionately excluded [32]. The high satisfaction levels among beneficiaries (75% reported being “quite” or “very satisfied”) indicates that existing schemes, though limited in reach, are perceived as valuable by those who can access them. Similar level of satisfactions were reported in Nigeria and Ethiopia among community members covered by a health insurance scheme [37,38].

Barriers to coverage were predominantly financial, with 37.1% of uninsured respondents mentioning high costs as the primary obstacle. Unawareness of insurance schemes (31.6%) and geographic constraints (7.1%) further compounded the problem, particularly in remote areas. These challenges call for targeted measures, such as subsidized subscription for the most disadvantaged social strata, public awareness campaigns, and the expansion of private health insurance including the promotion of community-based models to reach marginalized populations [14,33,39].

The study also revealed that health insurance significantly improved healthcare access and reduced perceived barriers. Insured individuals were more likely to use healthcare services (9.6%; *p* = 0.028) and reported fewer concerns about access (23.4%; *p* < 0.001). These results align with global evidence that health insurance mitigates financial hardships and encourages timely care-seeking behavior [40,41].

Most of the respondents (68%) expressed willingness to pay higher taxes for improved healthcare access, suggesting public support for systemic reforms. Our finding corroborates conclusions from studies conducted elsewhere in the world [5,42,43]. This finding could inform policy decisions, suggesting that tax-funded UHC schemes may be politically viable and socially desirable in Cameroon.

## Limitations

The cross-sectional design limits causal inferences, and self-reported data might introduce recall bias. However, the use of nationally representative sampling and robust statistical methods enhances the generalizability of the findings. The study’s focus on both coverage patterns and barriers provides a comprehensive foundation for future research and policy formulation.

## Conclusions

Cameroon’s suboptimal health insurance coverage reflects systemic inequities tied to employment, education, and geography. Addressing these gaps requires multi-factorial strategies, including expanding employer-independent schemes, subsidizing premiums for low-income groups, and leveraging community-based models. Policymakers should prioritize UHC initiatives to align with the population’s readiness for collective financing, ensuring equitable access to healthcare for all Cameroonians.

## Data Availability

All data produced are available online at : https://www.afrobarometer.org/data/data-sets/?select-countries%5B%5D=cameroon&hidden-current-page=1#listing

## Abbreviation

aOR: Adjusted odds ratio
CI: Confidence interval
cOR: Crude odds ratio
DALY: Disability-adjusted life year
GDP: Gross domestic production
OOP: Out-of-pocket payment
UHC: Universal health coverage

## Declaration

## Author contributions

FZLC conceived the original idea of the study, performed the analyses and wrote the first draft of the manuscript. FZLC, CTA, SD, GSN, JG-Y, AN, EOG, RT, EAE, AC, and IT critically reviewed and revised successive drafts of the manuscript. All authors read and approved the final manuscript.

## Ethical approval statement

Not applicable

## Consent for publication

Not applicable.

## Availability of data and materials

Data source of this study is available online in the Afrobarometer website [25].

