## Additional Files for "Healthcare coverage and associated factors in Cameroon: analyses from a national survey"

Supplementary Table 1 Medical coverage by subdivisions in Cameroon, 2024 (n = 1,200)

| **Division of residence** | **Medical coverage**  **n (%)** *^1^* | |
| --- | --- | --- |
|  | No  n = 1,112 | Yes  n = 88 |
| Bamboutos | 15 (93.8) | 1 (6.3) |
| Bénoué | 85 (96.6) | 3 (3.4) |
| Diamaré | 49 (87.5) | 7 (12.5) |
| Dja-et-Lobo | 8 (100.0) | 0 (0.0) |
| Fako | 49 (87.5) | 7 (12.5) |
| Faro | 8 (100.0) | 0 (0.0) |
| Haut-Nyong | 14 (87.5) | 2 (12.5) |
| Hauts-Plateaux | 23 (95.8) | 1 (4.2) |
| Kadey | 14 (87.5) | 2 (12.5) |
| Lekié | 39 (97.5) | 1 (2.5) |
| Lom-et-Djerem | 22 (91.7) | 2 (8.3) |
| Mayo-Banyo | 16 (100.0) | 0 (0.0) |
| Mayo-Danay | 37 (92.5) | 3 (7.5) |
| Mayo-Kani | 41 (85.4) | 7 (14.6) |
| Mayo-Louti | 16 (100.0) | 0 (0.0) |
| Mayo-Rey | 15 (93.8) | 1 (6.3) |
| Mayo-Sava | 15 (93.8) | 1 (6.3) |
| Mayo-Tsanaga | 20 (83.3) | 4 (16.7) |
| Mbam-et-Inoubou | 24 (100.0) | 0 (0.0) |
| Mbam-et-Kim | 14 (87.5) | 2 (12.5) |
| Mbéré | 8 (100.0) | 0 (0.0) |
| Mefou-et-Afamba | 13 (81.3) | 3 (18.8) |
| Mefou-et-Akono | 7 (87.5) | 1 (12.5) |
| Meme | 36 (90.0) | 4 (10.0) |
| Menoua | 7 (87.5) | 1 (12.5) |
| Mezam | 99 (95.2) | 5 (4.8) |
| Mfoundi | 122 (95.3) | 6 (4.7) |
| Mifi | 16 (100.0) | 0 (0.0) |
| Moungo | 27 (84.4) | 5 (15.6) |
| Mvila | 15 (93.8) | 1 (6.3) |
| Ndé | 7 (87.5) | 1 (12.5) |
| Nkam | 8 (100.0) | 0 (0.0) |
| Nyong-et-So'o | 16 (100.0) | 0 (0.0) |
| Océan | 15 (93.8) | 1 (6.3) |
| Sanaga-Maritime | 7 (87.5) | 1 (12.5) |
| Vina | 30 (93.8) | 2 (6.3) |
| Wouri | 155 (92.3) | 13 (7.7) |
| *^1^ p-value of the Fisher exact test = 0.430* | | |

Supplementary Table 2 Medical coverage by regions in Cameroon, 2024 (n = 1,200)

| **Region of residence** | **Medical coverage**  **n (%)** *^1^* | |
| --- | --- | --- |
|  | No  n = 1,112 | Yes  n = 88 |
| Adamawa | 54 (96.4) | 2 (3.6) |
| Centre | 235 (94.8) | 13 (5.2) |
| East | 50 (89.3) | 6 (10.7) |
| Far-North | 162 (88.0) | 22 (12.0) |
| Littoral | 197 (91.2) | 19 (8.8) |
| North | 124 (96.9) | 4 (3.1) |
| North-West | 99 (95.2) | 5 (4.8) |
| South | 38 (95.0) | 2 (5.0) |
| South-West | 85 (88.5) | 11 (11.5) |
| West | 68 (94.4) | 4 (5.6) |
| *^1^ p-value of the Fisher exact test = 0.041* | | |
